# A prediction algorithm to improve the accuracy of the Gold Standard Framework Surprise Question end-of-life prognostic categories in an acute hospital admission cohort-controlled study. The Proactive Risk-Based and Data-Driven Assessment of Patients at the End of Life (PRADA)

**DOI:** 10.1101/2023.09.07.23295196

**Authors:** Baldev Singh, Nisha Kumari-Dewat, Adam Ryder, Vijay Klaire, Gemma Bennion, Hannah Jennens, Dawn Matthews, Sophie Rayner, Benoit Ritzenthaler, Jean Shears, Kamran Ahmed, Mona Sidhu, Ananth Viswanath, Kate Warren, Emma Parry

## Abstract

**Objective:** To determine the accuracy of a clinical data algorithm allocated end-of-life prognosis amongst hospital inpatients.

**Method:** The model allocated a predicted Gold Standard Framework end-of-life prognosis to all acute medical patients admitted over a 2-year period. Mortality was determined at 1 year.

**Results:** Of 18,838 patients, end-of-life prognosis was unknown in 67.9%. A binary logistic regression model calculated 1-year mortality probability (*X^2^*=6650.2, p<0.001, r^2^ = 0.43). Probability cut off points were used to triage those with unknown prognosis using the GSF Surprise Question “Yes” or “No” survival categories (> or < 1 year respectively), with subsidiary classification of “No” to Green (months), Amber (weeks) or Red (days). This digitally driven prognosis allocation (100% vs baseline 32.1%) yielded cohorts of GSFSQ-Yes 15,264 (81%), GSFSQ-No Green 1,771 (9.4%) and GSFSQ-No Amber or Red 1,803 (9.6%).

There were 5,043 (26.8%) deaths at 1 year. In Cox’s survival, model allocated cohorts were discrete for mortality (GSFSQ-Yes 16.4% v GSFSQ-No 71.0% (p<0.001). For the GSFSQ-No classification, the mortality Odds Ratio was 12.4 (11.4 – 13.5) (p<0.001) vs GSFSQ-Yes (c-statistic of 0.71 (0.70 – 0.73), p<0.001; accuracy, positive and negative predictive values of 81.2%, 83.6%, 83.6% respectively. If this tool had been utilised at the time of admission, the potential to reduce subsequent hospital admissions, death-in-hospital, and bed days was all p<0.001.

**Conclusions:** The defined model successfully allocated end-of-life prognosis in cohorts of hospitalised patients with strong performance metrics for prospective 1 year mortality, yielding the potential to provide anticipatory care and improve outcomes.

**What is already known about this topic?:** End-of-life care is fragmented with excessive hospital admission and death in hospital. Current processes to determine end-of-life prognosis and promote anticipatory care for better outcomes are of limited utility.

**What this paper adds?:** A patient centric data integration model permitted the development of a digital health care system (PRADA) which allows the use of advanced analytics to accurately determine end-of-life prognosis among those where it was otherwise unknown. This paper demonstrates the potential benefit of integrating this prediction tool into routine care, at scale, in large population-level cohorts.

**Implications for practice, theory, or policy:** In an era of advancing opportunity from informatics driven heath care, NHS policy, through commissioning to direct care, must now actively deploy such evidence-based digital systems into direct care, most specifically in data sharing across provider boundaries. We particularly hope the research community might consider testing and validating this approach.

## Introduction

The NHS is under increasing pressure as the population ages and their needs become more complex, fragmented, and poorly integrated between service providers (1). End-of-life (EOL) is interposed by escalating rates of non-elective admissions (NEA) and death in hospital (DIH) (2–5). Guidance for better EOL care targets the reduction of NEA and DIH (6–14) from which there are emergent models of care (15). Hospital Specialist Palliative Care Teams (SPCT) improve EOL outcomes (16–17), but they may not be supported by effective prediction engines and clinical prognosis protocols (18,19). Specifically, the commended GSF Surprise Question (GSFSQ), establishing EOL prognosis, has shown variable performance (20,21). Current mortality prediction tools are not always automated and have uncertain performance characteristics (22–29), though the utilisation of modern-day informatics to support care provision through a population-based approach with early identification and systemised care is inherent to NHS digital health plans (30,31).

In our local health economy, we have integrated data from acute hospital, community and primary care sources and deployed it into direct care to promote early identification for anticipatory care, captured required process for structured care, developed linked clinical systems, and ascertained prospective events to determine effectiveness. We have termed the specific EOL digital care pathway; The Proactive Risk-Based and Data-Driven Assessment of Patients at the End of Life (PRADA). Our preliminary evidence has shown the potential benefit (32). Now we report a prediction model in which we have not attempted to use data prediction to usurp clinical judgment but rather to provide enhanced intelligence in support of targeted direct care and improved outcomes.

## Methods

### The PRADA systems

PRADA is an informatics driven digital health care system. It holds data, under full GDP compliance, of all patients registered to a Wolverhampton General Practice (GP), those resident in Wolverhampton but not registered to a local GP, and any who have had non-elective contact with our local hospital. Specific to the PRADA digital care pathway: risk escalators are applied to stratify the whole population followed by clinical stratification using the GSF Surprise Question which is captured at the point of care in hospital, community, and primary care settings. It ascertains the key EOL processes of EPaCCS, ReSPECT, EOL care plans, and EOL registration and key events including death, place of death, and non-elective urgent care. It has structured clinical governance functions and incorporates digital robotic processes in algorithms for clinical decision support such as highlighting individuals who may need to be on the EOL register. The PRADA clinical module is the deployment of that informatics resource to direct clinical care. It is accessed either through the electronic patient record (EPR) or as a standalone function. The system displays lists of patients in the specific arena of care that have been risk stratified as potentially requiring assessment for EOL care, so promoting proactive, anticipatory, and systematic care within non-referral pathways. It captures clinician data at the point of care, creates an EPaCC and a GSF prognosis specific EOL care plan, and automatically distributes finalised documentation electronically into the hospital EPR and to the GP patient record. At all times, the clinical module is linked to the core data system which then remains continuously up to date.

### Study design and setting

This is an acute hospital cohort-controlled study. Over 2-years, all who were admitted acutely as a non-elective admission (NEA) was ascertained. To assure a tighter case mix, only patients from the medical block of wards were included. The index admission (IA) was either the first admission in that period, or the one in which a PRADA assessment was first made, thereby excluding all recurrent or multiple admissions in any given individual, whilst yielding a time anchor for retrospective and prospective events.

### Data

The Wolverhampton Integrated Clinical Data Set links primary care, hospital, and community services data under GDPR. Demographic variables included age, gender, ethnicity, and the Index of Multiple Deprivation (IMD). Ethnicity data from all sources were recoded into White, South-Asian, Black, Mixed Ethnicity, Chinese or Unknown. The comorbidities utilized were the 16 commonest long-term conditions in the population. The variables chosen for digital risk stratification were based on common variables used in other risk prediction tools and assessed in our preliminary work to be linked to emergency activity and mortality. They represented urgent care and care complexity: >=3 Accident and Emergency (A&E) admissions over the prior 12 months (defined as emergency department attendances not leading to a hospital admission); >= 3 non-elective admissions (NEA) over the previous 12 months; >=3 co-morbidities, nursing home residency. The Rockwood score was used to assess frailty, or in its absence the Electronic Frailty Index, both categorised to moderate and severe frailty, whilst any missing vales coded to non-frail. Markers of end-of-life included end-of-life registration (EOL) or any recorded EOL process measure (EPaCC, ReSPECT or EOL care plans). The outcomes of the GSF Surprise question “Would you be surprised if this patient died within the next year?” was captured in the EPR together with the prognostic subcategories of “No” being Green (months), Amber (weeks) and Red (days). Any Specialist Palliative Care Team (SPCT) contact in the IA or in prior admissions was identified from routine inpatient coding. Mortality was determined from hospital mortality statistics and rolling NHS Strategic Tracing Service checks.

### Outcomes

Prospectively, death up to 1 year, and up to 90 days death in hospital, NEA and the number of hospital bed days occupied.

### Statistical method

All data were analysed on IBM SPPS version 29. When comparing independent groups, the Student’s t-test and the Chi-square test were used for the difference between means and proportions, respectively. Comparison of multiple group means was undertaken by one way analysis of variance (ANOVA). Variable taxonomy and collinearity were considered in principal components analysis (PCA). The association of independent factors with outcomes was achieved using binary logistic regression, presented with Odds Ratios (OR) and their 95% confidence intervals. Survival analysis was by Cox’s regression. Results are presented as the mean ± SD or as numbers with percentages. Statistical significance of all tests applied was taken at p<0.05.

### Research checklist

Strobe

### Ethical Considerations

The study did not involve randomisation or any intervention that ought not to have otherwise happened and so ethical approval was not deemed necessary, and this was confirmed by our authorised local governance group.

### Patient consent for publication

Not applicable.

### Patient and Public Involvement

None.

## Results

### Baseline characteristics

There were 142,159 admissions in those aged >=18 years old, with 91,244 individual single hospital spells, of which 18,838 were medical cases. Of those, 1,826 (9.7%) died in the index admission in hospital, 17,012 (90.3%) were discharged alive but 3,217 (17.1%) then died within 1 year of that discharge, totalling 5,043 (26.8%) deaths. Clinical characteristics are shown in **Table 1**. Those who died were older, more likely of white ethnic background, less deprived, had more A/E attendances and NEAs in the preceding year, were more co-morbid, more likely to be a nursing home resident, though they were equally frail, and were more likely on the end-of-life register and an end-of-life process. However, many of the differences were marginal and only 2 factors had > 50% association with death: >=3 NEA and EOL registration (53.9% and 65.5% respectively). The outcome of GSFSQ overwhelmingly shows the high prevalence with which it was not ascertained. Subsidiarily, there was a lower prevalence of GSFSQ-Yes and a higher prevalence of GSFSQ-No in those who died. **Table 1** also shows the Green (days), Amber (weeks) and Red (days) sub-prognostic classifications of the GSFSQ-No category. Among those with a GSFSQ-No classification or a hospital SPCT contact, 73.4% and 82.1% respectively died within 1 year. The SPCT saw 894 (65.7%) of patients with GSFSQ-No status (n=1,361).

**Table 1.** The characteristics of the cohorts. Data are the mean ± SD (range) of numbers with percentage. Abbreviations are as defined in the text.

|  | All | Alive at 1 year | Died at 1 year |  |
| --- | --- | --- | --- | --- |
| n | 18,838 | 13,797 (73.2%) | 5,043 (26.8%) |  |
| Age | 68.3±18.8 (18-110) | 64.4±19.3 | 78.9±12.4 | p<0.001 |
| Gender (male) | 9,483 (50.3%) | 6,891 (50.0%) | 2,592 (51.4%) | ns |
| Ethnicity (white) | 14,777 (78.4%) | 10,541 (76.4%) | 4,236 (84.0%) | p<0.001 |
| IMD | 27.6±16.3 (2.1 - 85.3) | 28.0±16.4 | 26.6±15.9 | p<0.001 |
| $\geq 3$AE attendances | 731 (3.9%) | 494 (3.6%) | 237 (4.7%) | p<0.001 |
| No. A/E attendances | 0.5 ±1.2 (0 - 45) | 0.45±1.22 | 0.53±1.05 | p<0.001 |
| $\geq 3$NEA | 345 (1.8%) | 159 (1.2%) | 186 (3.7%) | p<0.001 |
| No. NEA | 0.2±0.7 (0 - 20) | 0.12±0.59 | 0.32±0.96 | p<0.001 |
| NEA bed days prior | 9.7±16.9 | 6.4±14.0 | 18.8±20.5 | p<0.001 |
| Any comorbidity | 17,010 (90.3%) | 12,078 (87.6%) | 4,932 (97.8%) | p<0.001 |
| Total conditions | 3.2 ±2.2 (0 - 14) | 2.9±2.1 | 4.0±2.1 | p<0.001 |
| $\geq 3$ comorbidities | 10,664 (56.6%) | 6,918 (50.1%) | 3,746 (74.3%) | p<0.001 |
| Frailty (moderate or severe) | 5,319 (28.2%) | 3,624 (26.3%) | 1,695 (22.6%) | p<0.001 |
| NH resident | 1,825 (9.7%) | 932 (6.8%) | 893 (17.7%) | p<0.001 |
| EOL registration | 800 (4.2%) | 276 (2.0%) | 524 (10.4%) | p<0.001 |
| Any EOL process marker | 2,720 (14.4%) | 1,442 (10.5%) | 1,278 (25.3%) | p<0.001 |
| Seen by SPCT | 2,414 (12.8%) | 430 (3.1%) | 1,984 (39.8%) |  |
| Died in hospital in the IA |  |  | 1,826 (9.7%) (36.2%) |  |
| Died post discharge |  |  | 3,217 (17.1%) (63.8%) |  |
| Subsequent hospital death |  |  | 2,768 (14.7%) (54.9%) |  |
| Total hospital deaths |  |  | 4,594 (24.3%) (91%) |  |
| Days to death | | | 68.2±96.9 (0 - 365) | |
| Died at 30 days |  |  | 2,854 (15.7%) (56.6%) |  |
| Died at 90 days |  |  | 3,661 (19.4%) (72.6%) |  |
| **GSFSQ prognosis** |  |  |  |  |
| Unknown | 12,799 (67.9%) | 9,468 (68.6%) | 3,331 (66.1%) | p<0.001 |
| >1 year (GSFSQ -Yes) | 4,678 (24.8%) | 3,965 (28.7%) | 713 (14.1%) |  |
| <1 year (GSFSQ -No) | 1,361 (7.2%) | 362 (2.6%) | 999 (19.8%) |  |
| Green (months) | 530 (2.8%) | 222 (1.6%) | 308 (6.1%) |  |
| Amber (weeks) | 419 (2.2%) | 94 (0.7%) | 325 (6.4%) |  |
| Red (days) | 419 (2.2%) | 46 (0.3%) | 366 (7.3%) |  |

### Evaluation of variables

The variables listed in Table 1 were entered into a principal component analysis and redundant variables removed. At an Eigen value of >1, 4 distinct domains emerged explaining 41% of the data variance. These represented: clinical complexity (>=3 co morbidities, frailty, age, and nursing home residence); EOL (GSFSQ, SPCT contact, EOL process marker, EOL registration); Urgent care (>=3 NEA, >=3 AE attendances); and the residual demographics of IMD and ethnicity. Gender did not associate with any domain whilst age sat within the clinical complexity component. These variables were utilised in constructing a mortality prediction model.

### A regression model for mortality prediction

It is important to note that clinical judgment was used in the prediction model (any prior known recorded GSFSQ outcome) as well as being seen by the SPCT. The variables were entered into binary logistic regression with 1-year vital status as the dependent variable. The model was significant (*X^2^*=6650.2, p<0.001, r^2^ = 0.43), with a 46.6% prediction of those who died. It generated a predictive probability for death at 1 year which was ranked into quintiles. The 5^th^ quintile (predicted probability 0.7 ±0.17 (0.50 – 1.0)) stood out by having a 70.9% mortality rate capturing 53.0% of all deaths and in its association with the GSFSQ category (GSFSQ Unknown, Yes, No of 0.29±0.25, 0.18±0.18, 0.81±0.20 respectively (F=4090.3, p<0.001)).

### Triaging the GSF SQ

The 5^th^ quintile probability prediction was used to reclassify the GSFSQ. **Figure 1** shows a flow schematic of that process.

**Figure 1.**
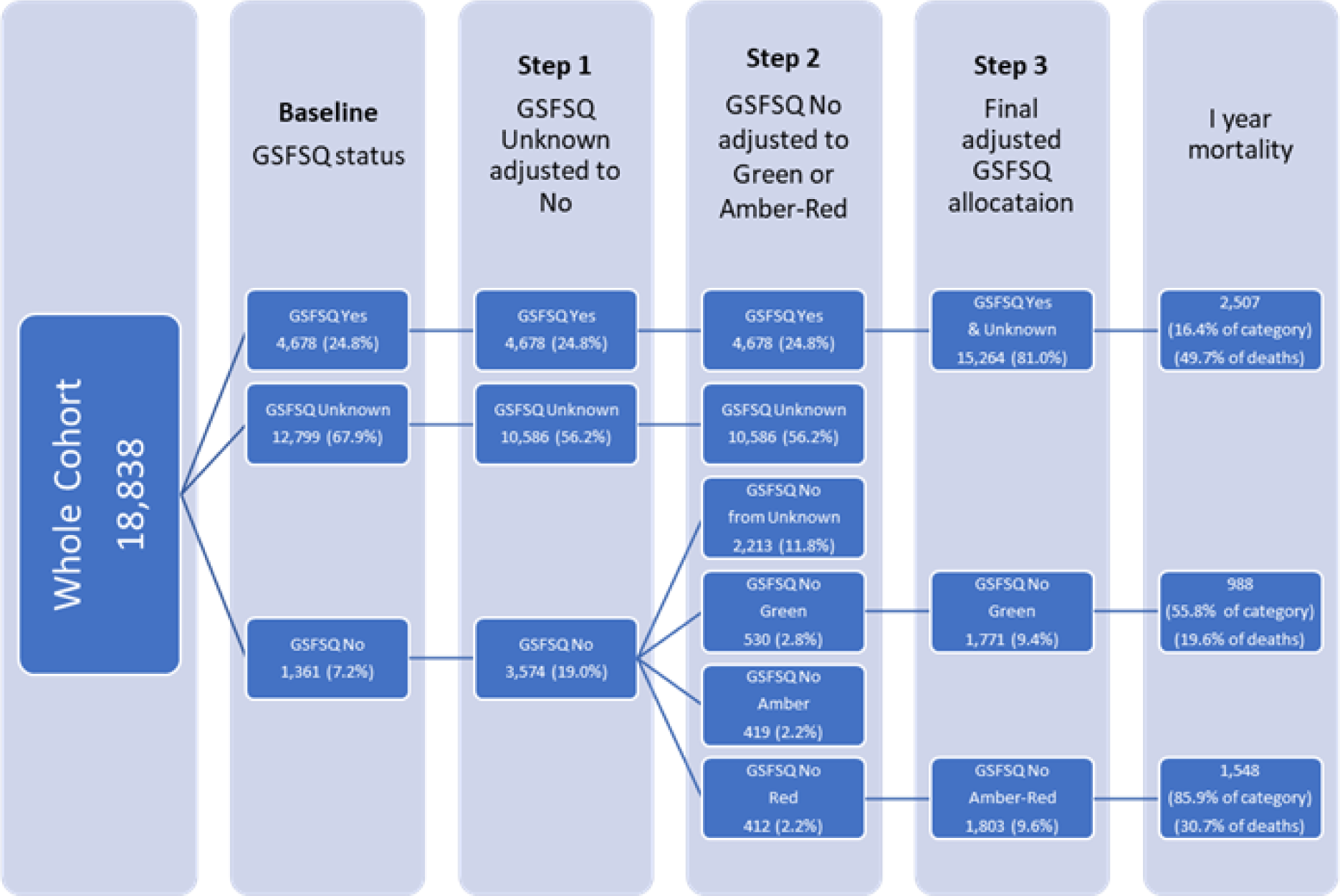
A flow diagram of the adjustments to the GSF SQ prognostic allocation made using the model’s probability estimates.

### Triage adjustment 1

Using the 5^th^ quintile category, in the first adjustment, GSFSQ-Unknown was reclassified to GSFSQ-No, reallocating 2,213 (17.3%) of the 12,799 with an unknown prognostic classification. The adjusted GSFSQ-No then captured 2,536 (50.3%) of those that died vs 999 (19.8%) prior to adjustment, meaning an additional 1,537 deaths (30.5% of all deaths). Reallocating the known GSFSQ-Yes group by 5^th^ quintile model predicted probability was of scant numerical value and was not incorporated.

**Figure 2.**
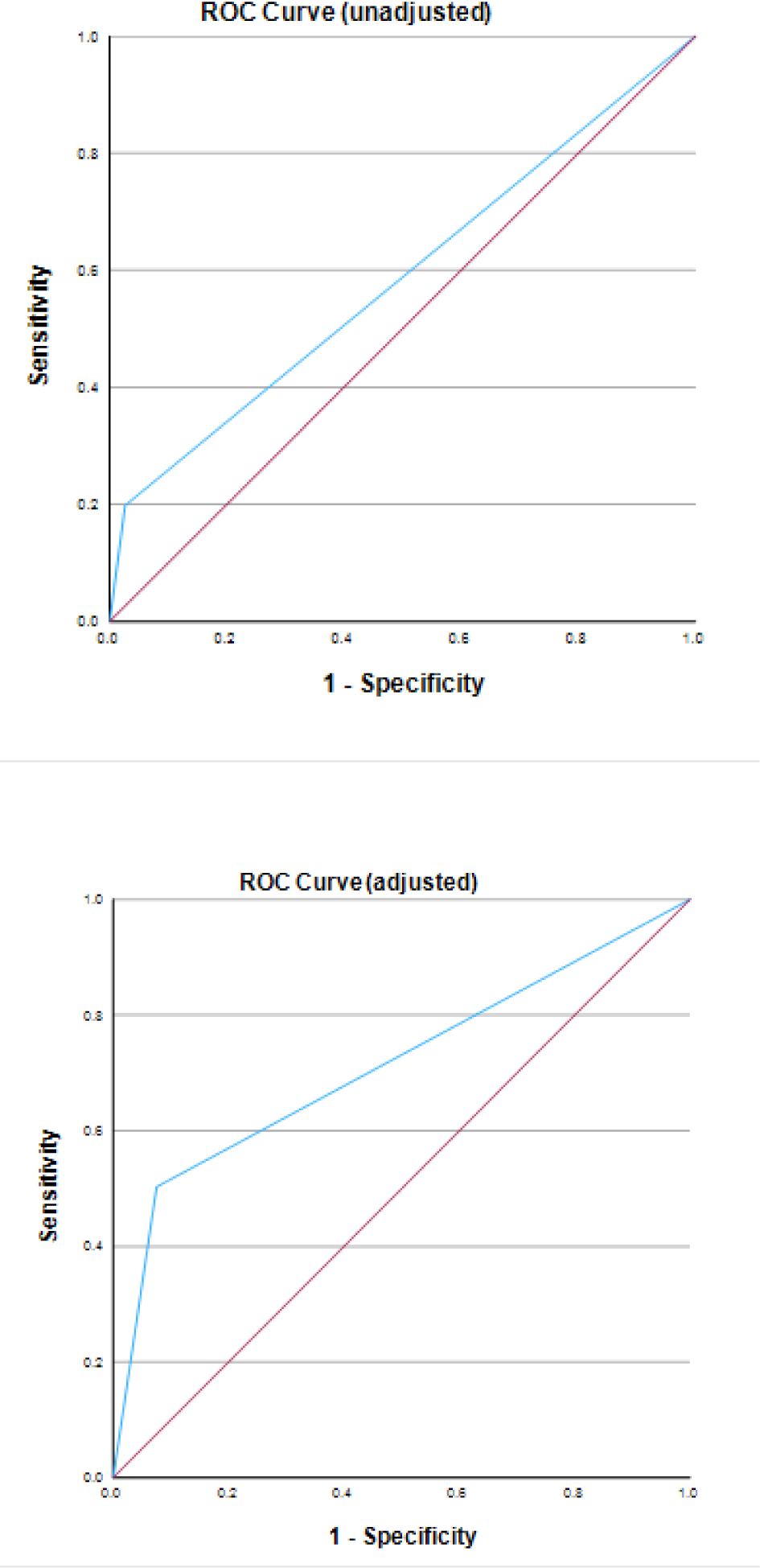
ROC curves assessing the GSFSQ-No category probability of death at 1 year either unadjusted (top) or adjusted by the described prediction model.

### Triage adjustment 2

The subsidiary prognosis for the GSFSQ-No was then further considered. In ANOVA (F=435.5, p<0.001), for the 5 baseline GSFSQ prognostic categories, GSFSQ-No Amber vs GSFSQ-No Red did not differ in probability scores in post hoc analysis, and they were merged as Amber-Red. The model probabilities for GSFSQ-N Green (0.76±0.21) and GSFSQ-No Amber-Red (0.86 ±0.18) were significantly different (p<0.001) with 95% CI for their means of 0.74 – 0.78 and 0.85 – 0.87, respectively. All reallocated GSFSQ-Unknows and GSFSQ-No Green were reclassified to Green or Amber-Red at a predicted mortality probability cut point of 0.85, the lower CI for the baseline GSFSQ-No Amber Red probability mean. As shown in **Table 2**, ultimately, predominately with re-classification of GSFSQ-Unknown, substantially more deaths were then captured by the finally adjusted GSFSQ-No classification (2,536 vs 999), with an increase in adjusted Green prognosis (988 vs 308) and a marked increase in adjusted Amber-Red (1,548 vs 691). Cox’s regression survival curves are shown in **Figure 3**, demonstrating that the residual post adjusted GSFSQ-Unknown group then approximates very closely to the original GSFSQ-Yes, with preservation of the distinction between Green and Amber-Red.

**Figure 3.**
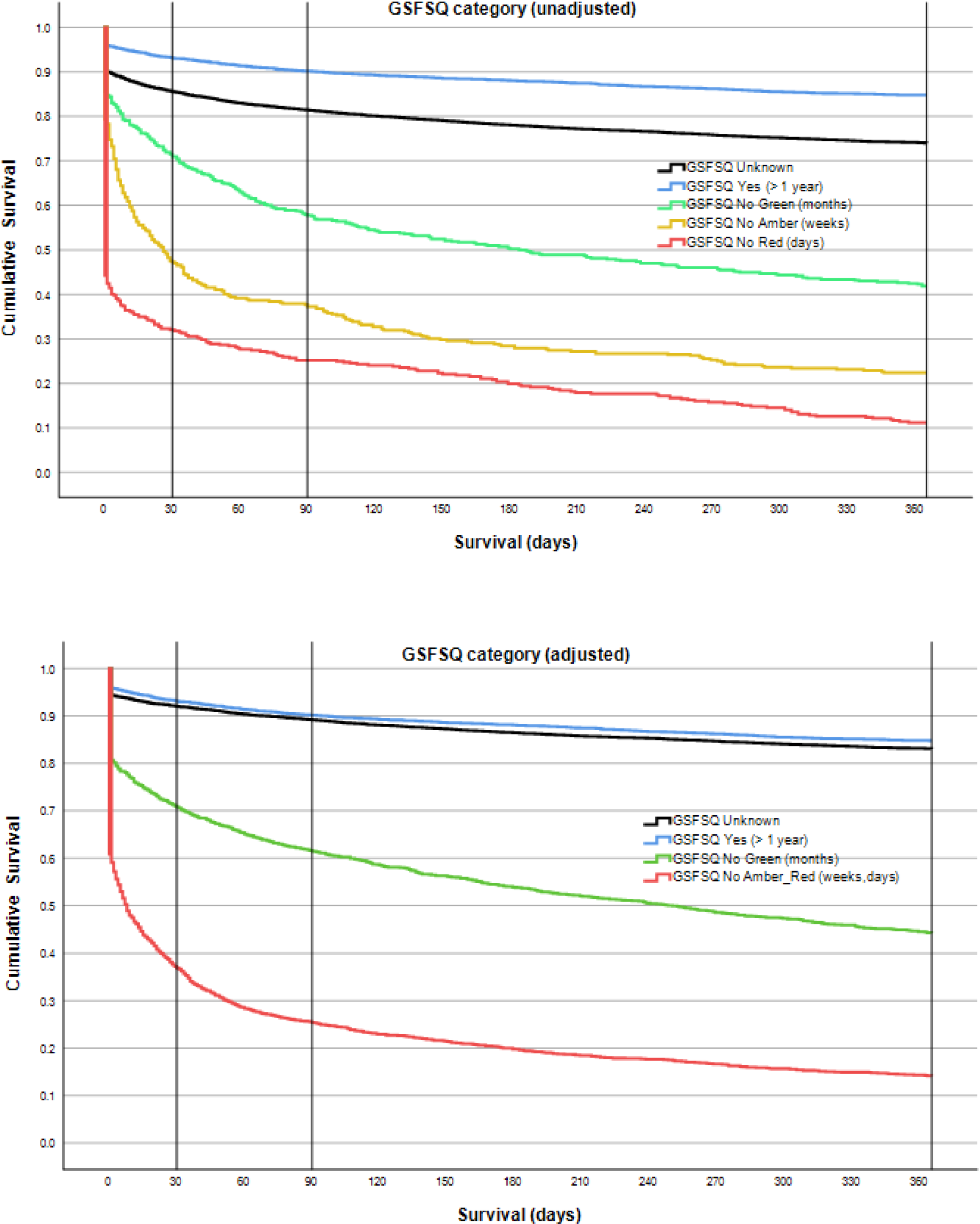
Survival curves for the GSFSQ categories in the unadjusted and adjusted model.

**Table 2.**
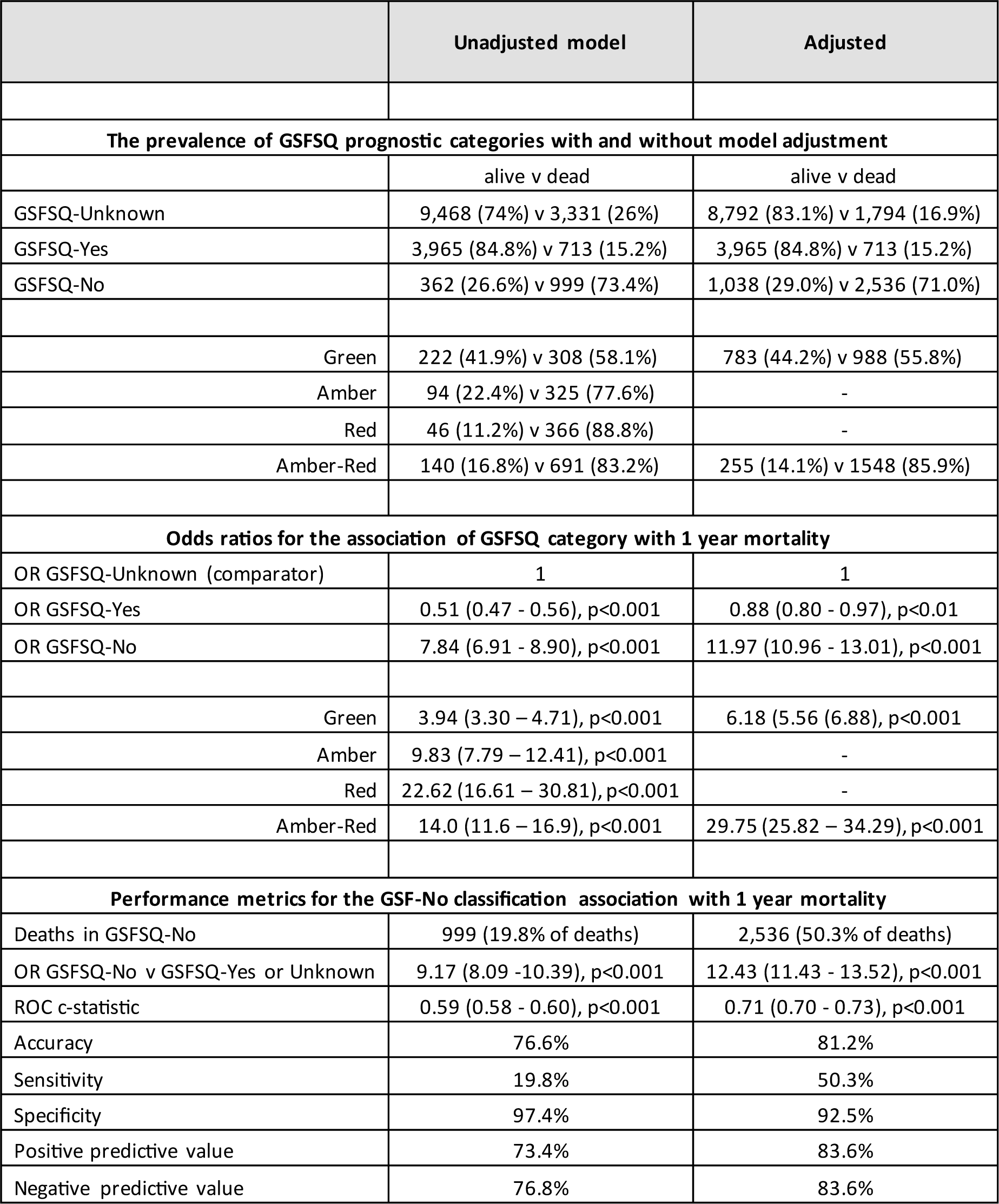
Model metrics with and without predicted probability adjustment. Odds ratios were determined in binary logistic regression and performance metrics were derived from ROC analysis.

|  | Unadjusted model | Adjusted |
| --- | --- | --- |
| **The prevalence of GSFSQ prognostic categories with and without model adjustment** |  |  |
|  | alive v dead | alive v dead |
| GSFSQ-Unknown | 9,468 (74%) v 3,331 (26%) | 8,792 (83.1%) v 1,794 (16.9%) |
| GSFSQ-Yes | 3,965 (84.8%) v 713 (15.2%) | 3,965 (84.8%) v 713 (15.2%) |
| GSFSQ-No | 362 (26.6%) v 999 (73.4%) | 1,038 (29.0%) v 2,536 (71.0%) |
| Green | 222 (41.9%) v 308 (58.1%) | 783 (44.2%) v 988 (55.8%) |
| Amber | 94 (22.4%) v 325 (77.6%) | - |
| Red | 46 (11.2%) v 366 (88.8%) | - |
| Amber-Red | 140 (16.8%) v 691 (83.2%) | 255 (14.1%) v 1548 (85.9%) |
| **Odds ratios for the association of GSFSQ category with 1 year mortality** |  |  |
| OR GSFSQ-Unknown (comparator) | 1 | 1 |
| OR GSFSQ-Yes | 0.51 (0.47 - 0.56), p<0.001 | 0.88 (0.80 - 0.97), p<0.01 |
| OR GSFSQ-No | 7.84 (6.91 - 8.90), p<0.001 | 11.97 (10.96 - 13.01), p<0.001 |
| Green | 3.94 (3.30 – 4.71), p<0.001 | 6.18 (5.56 (6.88), p<0.001 |
| Amber | 9.83 (7.79 – 12.41), p<0.001 | - |
| Red | 22.62 (16.61 – 30.81), p<0.001 | - |
| Amber-Red | 14.0 (11.6 – 16.9), p<0.001 | 29.75 (25.82 – 34.29), p<0.001 |
| **Performance metrics for the GSF-No classification association with 1 year mortality** |  |  |
| Deaths in GSFSQ-No | 999 (19.8% of deaths) | 2,536 (50.3% of deaths) |
| OR GSFSQ-No v GSFSQ-Yes or Unknown | 9.17 (8.09 -10.39), p<0.001 | 12.43 (11.43 - 13.52), p<0.001 |
| ROC c-statistic | 0.59 (0.58 - 0.60), p<0.001 | 0.71 (0.70 - 0.73), p<0.001 |
| Accuracy | 76.6% | 81.2% |
| Sensitivity | 19.8% | 50.3% |
| Specificity | 97.4% | 92.5% |
| Positive predictive value | 73.4% | 83.6% |
| Negative predictive value | 76.8% | 83.6% |

### Triage adjustment 3

Whilst the final adjusted GSFSQ-Yes still retained negative predictive value compared to final residual GSFSQ-Unknown (OR 0.88 (0.80 - 0.97), p<0.01), the 2 could be merged, so that the final outcomes were essentially GSFSQ-Yes (15264 (81.0%)), GSFSQ-No Green (1771 (9.4%)) and GSFSQ-No Amber-Red (1803 (9.6%)) encapsulating 2,507 (49.7%), 988 (19.6%) and 1,548 (30.7%) of deaths, respectively.

### Model performance metrics

These are shown in **Table 2** with ROC curves in **Figure 2**. The model adjusted GSFSQ No captured 50.3% of deaths, its OR for predicting death was stronger, the ROC statistic improved, as did accuracy and positive and negative predictive values.

### Workload considerations

The purpose of this analytical strategy was not to predict mortality but drive accurate early identification of those at risk of dying, targeting them for systematically delivered assessment and interventions that might lead to benefit in better outcomes. Regarding workload, ***Table 3*** shows the association between GSFSQ status and palliative care team actual and potential input requirements. Regrading of the GSFSQ categories within the adjusted model, this demonstrates a truer reflection of the actual appropriateness of SPCT contact, which was focused in the highest risk categories and progressively less associated with the inaccurate baseline GSFSQ-Unknown status. As greater numbers are identified, the table shows how SPCT workload might change if the SPCT team then saw various categories of patients, including estimates of daily new caseload. In the adjusted model, seeing all identified GSFSQ-No patients would increase workload by 52%, restricting to only seeing those with GSFSQ-No Amber-Red status reduced it by 24% whilst seeing all GSFSQ-No Amber-Red additional to current caseload, increased it by only 10%.

### Estimations of benefit

Estimating benefit in hard outcomes is difficult to determine and the analysis is hypothetical. Using the described recently deployed PRADA systematic care planning tool (32), where numbers remain small in the context of this 2-year cohort, we selected those discharged alive and in the highest propensity adjusted GSFSQ-No Amber-Red category. The associations with 90-day post discharge events are shown in **Table 3**. There were 1,102 such cases of whom 123 (11.2%) had a PRADA digitally supported EOL care planning contact with the SPCT. The PRADA contact group had a higher mortality rate but nevertheless had fewer hospital admissions, occupied less hospital bed days and were less likely to die in hospital (all p<0.001). The potential emergency bed day saving over 90-days was 3.53 days per non-PRADA contact patient. This equates to a total of £1,727,935 (979 x 3.53 x £500) or £1, 765 per individual. The combined adverse hard outcome event of 90-day NEA or hospital death was also significantly lower in the PRADA group (p<0.001). For PRADA contact the relative risk of this summative adverse event was 0.51 (0.35 – 0.73) (p<0.001) and the NNT for that benefit were 5.2 (3.5-9.7).

**Table 3.** The association between current and modelled GSFSQ status and the specialist palliative care team (SPCT) current and potential activity, and an estimate of potential benefit of contact when seen using the PRADA system.

|  | **Activity** |  |
| --- | --- | --- |
|  | **Not seen by the SPCT v Seen (n=2, 414) with row and column %** |  |
|  | Unadjusted | Adjusted |
| GSF Unknown | 11,561 v 1,238 (9.7%) (51.3%) | 10,385 v 201 (1.9%) (8.3%) |
| GSF Yes | 4,396 v 282 (6.0%) (11.7%) | 436 v 282 (6.0%) (11.7%) |
| GSF No | 467 v 894 (65.7%) (37.0%) | 1,643 v 1,931 (54.0%) (80.0%) |
| Green | 310 v 220 (41.5%) (9.1%) | 1,408 v 363 (20.5%) (15.0%) |
| Amber or Red | 157 v 674 (81.1%) (27.9%) | 235 v 1,568 (87.0%) (65.0%) |
|  | **Potential activity (adjusted model)** |  |
| Total currently seen by SPCT | 2,414 (current daily caseload 2.8) |  |
| Caseload if only all GSFSQ-No seen | 3,547 (+1,133) (estimated daily caseload 4.2 (+1.4, +50%)) |  |
| Caseload if only all GSFSQ-No Amber or Red seen | 1,803 (-611) (estimated daily caseload 2.1 (-0.7, -25%)) |  |
| Caseload if all GSFSQ-no Amber or Red are additional | 2,649 (+235) (estimated daily caseload 3.1 (+0.3, +10%)) |  |
|  | **90-day Outcomes amongst those discharged alive with GSFSQ-No Amber Red status (adjusted model, n= 1,102), not seen in PRADA based contact vs seen.** |  |
| Non-PRADA vs PRADA contact group | 979 (89%) v 123 (11%) |  |
| 90-day death | 559 (57.1%) v 88 (71.5%), p<0.01 |  |
| 90-day NEA | 378 (38.6%) v 23 (18.7%), p<0.001 |  |
| 90-day hospital bed days | 4.64±10.4 v 1.11± 3.8, p<0.001 |  |
| Died in hospital | 143 (25.6%) v 4 (4.5%), p<0.001 |  |
| 90-day NEA OR died in hospital | 380 (38.8%) v 24 (19.5%), p<0.001 |  |

## Discussion

### Main findings

The model’s algorithm allowed end-of-life prognosis allocation, in line with the GSF SQ, amongst those who had not already been given a prognosis by clinicians, characterising the whole cohort, in manner that had strong predictive accuracy for 1 year mortality outcomes. Running this algorithm in live clinical care has the potential for anticipatory care by a SPCT to modify key outcomes leading to reduced rates of non-elective readmission, bed occupancy, and in hospital mortality.

### What this study adds

The study defines how to take whole health economy clinical data and curate a limited data set to run an automated predictive model. The methodology for probability cut off points that characterise the intended prognostic groups are outlined, and the accuracy of their association with various relevant endpoints, most especially 1 year mortality, is demonstrated.

### Strengths and Weaknesses

The study’s strengths are the use of said integrated clinical data set, the construction of a digital heath care pathway specific to the end-of-life, the use of it to run advanced analytics and the description of that process in addition to the ascertainment of the performance metrics of its association with 1 year mortality. The findings are likely generalisable to other such care settings given the large size of the cohort of a general medical case mix, not limited to small numbers, nor in niche clinical scenarios. Its weakness is that the predicted end-point benefits were theoretical and now require evidencing in real time care, although we have published our positive preliminary outcomes (32). Furthermore, the integrated clinical data between acute, community and primary care provision is not readily available in most health economies. Finally, as a first report, the findings require validation, in various care settings care, by independent research groups. The study relates to those with a hospital admission, and it is not yet known if the algorithm might apply to the non-admitted population.

### General discussion

We describe the model not as predicting mortality but predicting a possible clinical decision in a form equivalent to the GSFSQ. This may be a moot point, they may be seen as the same thing, but our prediction methodology is intended to support clinical decision making and not to undermine or replace it in any way and it would be ethically very unsound to assume that the model predicated GSFSQ is the actual prognosis or for any care process to run automatically on that basis. Thus, in our model, established clinical judgment remains the prime decision, and the predicted output is simply an effective risk-stratification tool to drive further assessment by clinical teams for validation of the proposed digital allocation and for the planning of effective and appropriate care, ensuring it to be matched to the patient’s clinical needs, consent and wishes, which cannot be known to an automated prediction engine.

Much is made of the intuitive, ad-hoc, “gut feeling” of global clinical judgment (GCJ) (33–36), but here we incorporate it into probability model in line with a Bayesian approach to clinical decision making (37). We have recently published the positive power of the use of GCJ in a similar prediction engine (38). In end-of-life care there is already an established format for capturing GCJ, namely the GSF SQ although it has been criticized in several ways (20,21). However, our view is that: the overwhelming problem resides with the question not being asked; when it is asked its performance should not be judged as a discreet variable (as it has been hitherto) which is bound to be poor, just as with each other single independent variable (e.g. comorbidity); it is strengthened by incorporating it alongside other key relevant variables; it is statistically a valuable parameter as determined by principal component analysis. Furthermore, the focus should not always be on predicting who might die but clarifying who is unlikely to, by valuing its negative predictive value, a notion that resonate with clinicians, who may find it much easier to rule something out as opposed to ruling it in as time bound event as crucial as death (39,40). Thus, the final GSFSQ-Yes category excluded 81% of the cohort who had a much lower 1-year mortality of 16.4% and risk escalated 19% to the GSF-No category with a 70.1% mortality rate and achieving this level of risk stratification or “prediction” was the intended prime outcome. It maybe, but unknown to us at this stage, that further clinical validation, assessments and events would have again modified the prediction probabilities, emphasising that this is intended to be a live and continuous process, not a single time-point cross-sectional snapshot, and so probability adjustment can become dynamic with trends that might be informative.

Crucially, the models’ prognostic allocation is integrated in a digital care pathway and is used to drive anticipatory care by automated referral to the SPCT. The system will know if that SPCT had contact with the identified patients, whether required processes of care were undertaken, whether onwards care allocation occurred appropriately with effective communication to the relevant providers of care, especially community and primary care services. Prospective monitoring allows the ascertainment of the defined outcomes as well as an understanding end-of-life registration status, perhaps the most crucial event in the pathway. Thus, the next step of our work is to integrate the now defined analytical process into live routine care and to determine its impact across the whole PRADA digital pathway construct from identification to registration and the prevention of the adverse outcomes of escalating non-elective care and death not in the preferred place of death.

Briefly, PRADA is distinctly different from many other mortality prediction tools (22–29). They share an aspiration for early identification and anticipatory care. They may utilise the GSFSQ as a preliminary identification step. They rely on a variety of individual functional assessments and specific clinical categories or markers of end organ failure and these generally require summation by an individual clinician case by case. Accordingly, contrary to their ambition, they may be deployed reactively late in this trajectory and not proactively. Though they may have viable performance metrics, these have generally been assessed in small and specific groups of patients. Their algorithms are not generally automated into clinical systems and thus are not easily made systematic, uniform, deployable at scale or amenable to continuous quality improvement. However, PRADA methodology, whilst differing in the data sets utilised, shares the common principles of digitised risk stratification, the capturing of validating clinical assessment, prognostic coding, care planning and registration with the innovative NHS EARLY toolkit which can run in a similarly automated manner, directly in primary care systems to highlight specific patient in these end-of-life cohorts (29).

In summary, as part of a wider piece of work creating an end-of-life digital care pathway, PRADA, we have developed an informatics-based algorithm for prognosis prediction that maps to the GSFSQ categories and is validated to have performance accuracy against 1 year mortality. It can be run at scale in large cohorts of patients, in this case those who have had hospital admission. The next phase of our work is to demonstrate this can be automated live into direct care in our electronic clinical systems and demonstrate the hoped for consequent beneficial outcomes are achieved. Our work and methodology will have resonance not only in the developing UK NHS digital heath care ambition, but in many other health economies across the world faced with exactly similar imperatives. We would implore colleagues in the research community to consider and test our findings.

## Data Availability

All data produced in the present study are available upon reasonable request to the authors

## Author contributions

Principal author BMS; Authors EP, NK, AR, AV; Data systems VK; Statistical Analysis BMS; Manuscript Development All; PRADA system development and pilot AV, DM, GB, HJ, JS, KA MS, SR.

## Conflicts of Interest or Competing Interest

None

## Data Sharing

The corresponding author will share anonymised study data upon reasonable request.

## Copyright

The corresponding author has the right to grant on behalf of all authors, the license to permit this article to be published.

## Abbreviations Used

CI: confidence interval
DIH: death in hospital
EOL: end of life
EPaCCS: Electronic Palliative Care Co-ordination Systems
EPR: electronic patient record
GDPR: General Data Protection Regulation
GSF SQ: Gold Standard Framework Surprise Question
IA: index admission
IMD: Index of Multiple Deprivation.
NEA: non-elective admission
NHS: National Health Service
OR: Odd’s Ratio
PCA: Principal components analysis
PRADA: Proactive Risk-Based and Data-Driven Assessment of Patients at the End of Life
RCT: randomised controlled trial
ReSPECT: Recommended Summary Plan for Emergency Care and Treatment
ROC: receiver operating curve
SD: standard deviation
SPCT: specialist palliative care team

